# OncoVR - Virtual Reality in Oncology for Patient-centered Care: A Systematic Review and Meta-Analysis

**DOI:** 10.1101/2025.10.28.25338526

**Authors:** Miriam Balzer, Abdelrahman Elsakka, Gijs Luijten, Katrin Schormann, Slobodan Jovic, Monja Gerigk, Laura Mazilescu, Sophia Schmitz, Julius Keyl, Stefan Liszio, Oliver Basu, Beata Maria Goraus, Stefan Kasper, Jens Kleesiek, Behrus Hinrichs-Puladi, Jan Egger

## Abstract

Due to the significant research effort in the field of virtual reality (VR) and the increasing interest of the gaming industry, the technical possibilities have developed rapidly. In recent years, VR applications have become more extensive, the devices more convenient and less complex to handle. As a result, the benefits of VR have long since arrived in the clinical field and are being used in various areas. However, since VR has not yet made it into everyday use, average patients are not experienced in using these devices. In the past, VR applications for oncology patients have already been used to improve well-being and reduce stress, anxiety and nausea during therapy, surgery and diagnostic procedures. Oncology is particularly well suited to psychological VR applications because cancer can be a potentially traumatic experience that has a strong impact on the patient’s mental state, and because the mental state can also have a strong influence on the success of the therapy. In addition, oncological therapies are usually a long-term treatment, which is why studies on the extensive use of VR are a suitable option. This systematic review summarizes the current state of research in this area and discusses the possibilities for using VR in patient-centered care in oncology.

## 2 Introduction

Cancer is one of the most common diseases and a leading cause of death worldwide [1]. Despite significant advancements in cancer therapy, which have led to a substantial increase in survival rates for many forms of cancer, numerous therapeutic approaches continue to be associated with substantial side effects and limitations for patients. In addition to the well-researched physical side effects, such as hair loss, nausea, fatigue and infertility, it is primarily mental stress that affects cancer patients. This, for example, results in an increased risk of depression and anxiety [2]. Cancer therapies often have a major impact on the patient’s quality of life [3]. At the same time, the impairment of quality of life has proven to be a relevant factor for the life expectancy of cancer patients [4]. The fact that more cancer patients live longer as a result of better therapies also increases the number of traumatized patients and those who require psychotherapeutic support. However, we already have a massive shortage of psychotherapists (in Germany) with extremely long waiting lists [5]. It is therefore necessary to do everything possible to reduce the psychological burden on cancer patients. Ensuring the well-being of patients during therapy is therefore an essential contribution to the overall outcome of treatment and should always be incorporated into therapy. Psychological interventions, self-help groups, music and sports therapy and other similar strategies have been integrated into the treatment of cancer patients for some time now and are increasingly becoming routine [6, 7]. Drugs to relieve pain and reduce nausea and fatigue are also an integral part of cancer treatments. However, limiting factors are still the low number of psychological interventions available due to staff shortages and a lack of funding, as well as the side effects of the various drugs used to treat the side symptoms of therapy. Digital services with a lower inhibition threshold and fewer side effects are therefore a very desirable approach. Virtual reality (VR) is proving to be a promising tool in this regard. The concept of VR has its origins in Morton’s creation of the Telesphere Mask and the Sensorama from 1960 [8], and the theory of the ultimate display by Ivan Sutherland [9], but for a very long time it was considered an expensive and technically complex gimmick. This only changed with the founding of the first commercial VR company ‘VPL Research’ by Jaron Lanier in 1984 and further with the development of the first prototype of the Oculus Rift DK1 by the former manufacturer Oculus in 2012/2013 [10]. Even though it took a long time for VR to get implemented in daily practice, the first studies on the use of VR as a therapeutic tool were conducted as early as 1996 [11]. VR immerses users in simulated 3D environments, offering potential benefits in pain management, rehabilitation, and psychological support [12, 13]. VR can be divided into immersive and non-immersive VR. Non-immersive VR uses a combination of screens surrounding the user to display virtual information. One example of this is the CAVE system developed in the early 1990s [14]. In this way, information can be displayed in such a way that the user is at the centre of the action. Flight simulators are an example of this. Immersive VR refers to the use of a head-mounted display (HMD) to track a user’s movement and display VR information based on the user’s position, which intensifies the VR experience for the user [15]. This review will focus on the use of immersive VR. VR is an effective technology to distract people from pain and discomfort by immersing them in stimulating and calming environments, which can help to improve mood and well-being. Especially for cancer patients who often struggle with side effects such as pain and depression and who are often limited in their ability to distract themselves, VR offers a good alternative. Moreover, integrating VR into psychotherapy sessions offers a supportive environment for patients to cope with the emotional and psychological challenges of their disease. As technology continues to evolve, VR holds promise for further enhancing the quality of care and overall well-being of cancer patients undergoing treatment. This systematic review examines the potential of VR to improve the well-being of cancer patients during various forms of therapy to treat cancer. These include chemotherapy, radiotherapy and surgical interventions.

## 3 Virtual Reality Reviews in Oncology

### 3.1 Search Strategy

To ensure a systematic approach, this review was registered with PROSPERO under the ID CRD42024495611. The search strings as well as the inclusion and exclusion criteria were defined in detail. The aim was to collect all the literature in which VR was used in cancer patients during therapy (see Table 1). Three databases were selected for the systematic search, and the appropriate search strings for each database were defined. Reports dealing with both VR and oncology were queried. The different forms of therapy were also included in the search queries. The details of the queries can be found in Table 1. The query was performed on January 9, 2024 and resulted in 3264 reports.

**Table 1:** Table of the used databases and search strings together with the resulting number of reports for each query.

| Database | Search String | Records |
| --- | --- | --- |
| Pubmed | ("oncology"[Title/Abstract] OR "cancer"[Title/Abstract] OR "radiation*" [Title/Abstract] OR "radiotherap*" [Title/Abstract] OR "chemo*" [Title/Abstract] OR "Neoplasms"[MeSH Terms]) AND ("virtual eality"[Title/Abstract] OR "Virtual Reality"[MeSH Terms]) | 797 |
| Scopus | (TITLE-ABS(oncology) OR TITLE-ABS(cancer) OR TITLE-ABS(radiation*) OR TITLE-ABS(radiotherap*) OR TITLE-ABS(chemo*) OR TITLE-ABS(neoplasms)) AND(TITLE-ABS(virtual reality)) | 1152 |
| Web of Science | (TS=(oncology) OR TS=(cancer) OR TS=(radiation*) OR TS=(radiotherap*) OR TS=(chemo*) OR TS=(neoplasms)) AND TS=(virtual reality) | 1315 |

Only studies with original research conducted entirely by the authors were included. The studies had to be peer reviewed and focused primarily on the use of VR in conjunction with conventional cancer treatments to reduce potential side effects for patients. Studies with non-English full texts were excluded. Studies that were not exclusively original research studies, e.g., reviews or book chapters, were excluded. Studies without peer review, e.g., conference posters/abstracts, preprints, or commentaries, were also excluded. Studies that did not focus primarily on the use of VR alongside conventional cancer treatments to reduce potential side effects for patients, or where VR was not used by the patients themselves, were also excluded. The resulting reports were filtered for duplicates and then checked twice in a double-blind process using Rayyan [16]. In the first round, inclusion and exclusion criteria were applied to the titles and abstracts of the reports. Reports with titles and abstracts that clearly indicated that the publication did not fit the desired scheme were rejected. Both reviewers had to work independently and consult each other at the end if there were disagreements. If no agreement could be reached, an independent third consultant was appointed to make a final decision. The same procedure was used for the 2nd round, but here the full text of the publications was checked. Figure 1 shows the different stages of the review and the number of reports excluded at each stage.

**Figure 1:**
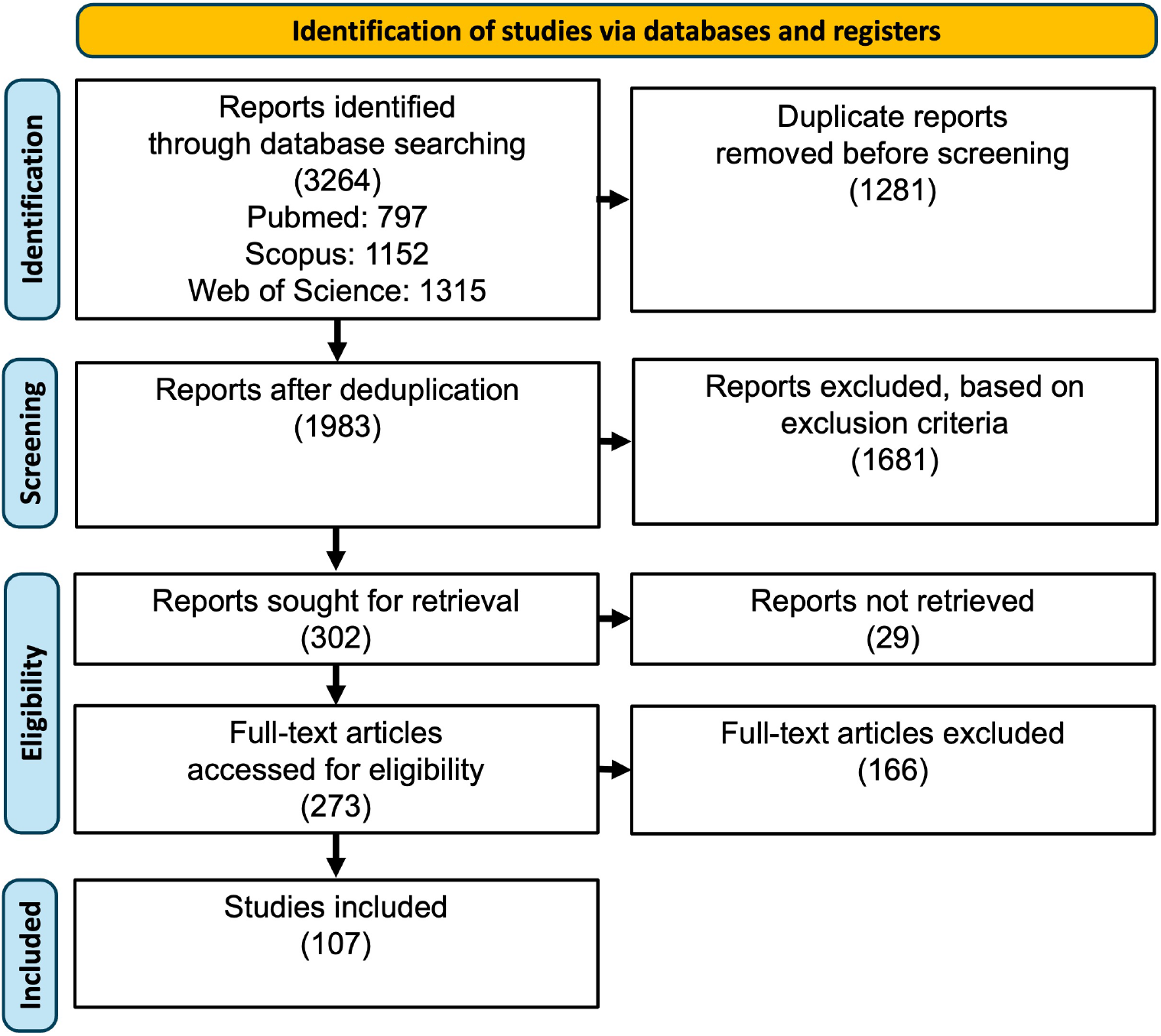
Search strategy used in this systematic review. Source: Adapted from the PRISMA flow diagram by Moher et al [17]. After removing duplicates and reports without available fulltext, a total of 273 reports were screened after the named exclusion criteria, resulting in a total of 107 cited reports.

### 3.2 Extracted Reviews

The procedure resulted in 107 included full papers for data extraction, which were then read in detail and sorted into categories. A total of nine categories emerged, which can be understood as a collective term for the corresponding reports: children, during chemotherapy, before therapy, during cancer treatment in general, during a surgical/invasive procedure, rehabilitation after a procedure, detection of side effects and palliative care. The subdivision was worked out step by step, firstly according to the age of the target group, then according to the timing of the VR application in the course of the therapy, then according to the goal of the VR application and finally according to the type of cancer therapy applied. Figure 2 explains the categorisation in detail.

**Figure 2:**
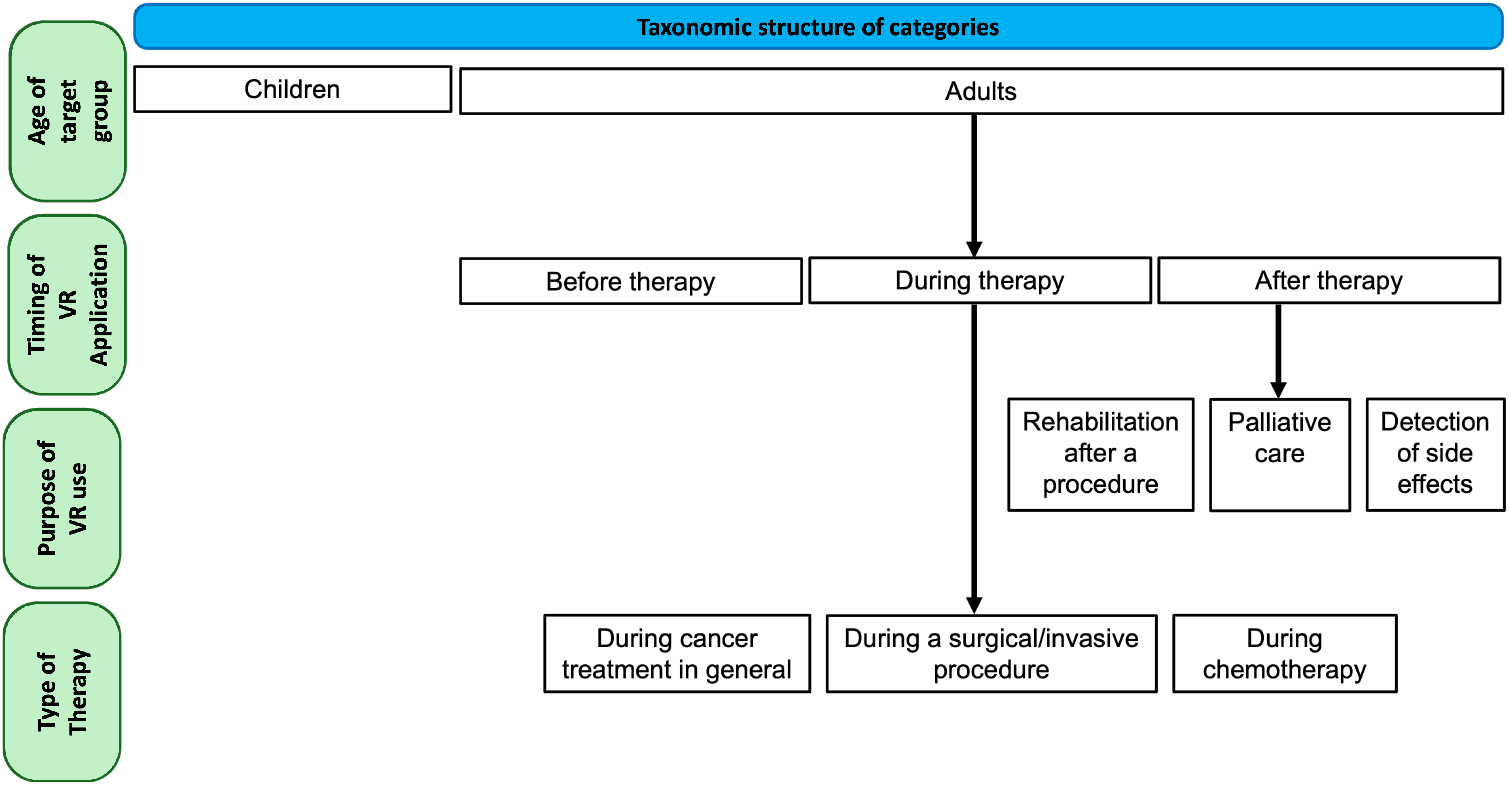
The criteria according to which the progressive subdivision into the various categories was carried out are shown.

The aim of the taxonomy into different subgroups was to generate similarly sized homogeneous groups for a better overview, within which comparability is ensured. Table 2 shows how many reports are part of each group.

**Table 2:** Number of reviews for each category.

| Group | Reports | References |
| --- | --- | --- |
| Children | 29 | [18–47] |
| During cancer treatment in general | 23 | [48–70] |
| During chemotherapy | 15 | [71–85] |
| Before therapy | 13 | [86–98] |
| During a surgical/invasive procedure | 11 | [99–109] |
| Palliative care | 8 | [110–117] |
| Rehabilitation after a procedure | 5 | [118–122] |
| Detection of side effects | 3 | [123–125] |

The categorization according to the specific area of application was much easier than the subdivision according to patient characteristics such as gender, age, or disease. Most of the studies were conducted on heterogeneous groups. However, in order to ensure an overview of the groups of people, a subdivision according to the type of cancer was made in Figure 3. Many reports are represented more than once in this figure, which is due to the heterogeneous group of study participants. This means that patients with different types of cancer were included in the respective study. In other reports, however, no information was provided on the type of cancer. This was often the case in palliative studies, for example, in which only terminal cancer was mentioned as a disease. Figure 3 illustrates the distribution of the reports across the various types of cancer.

**Figure 3:**
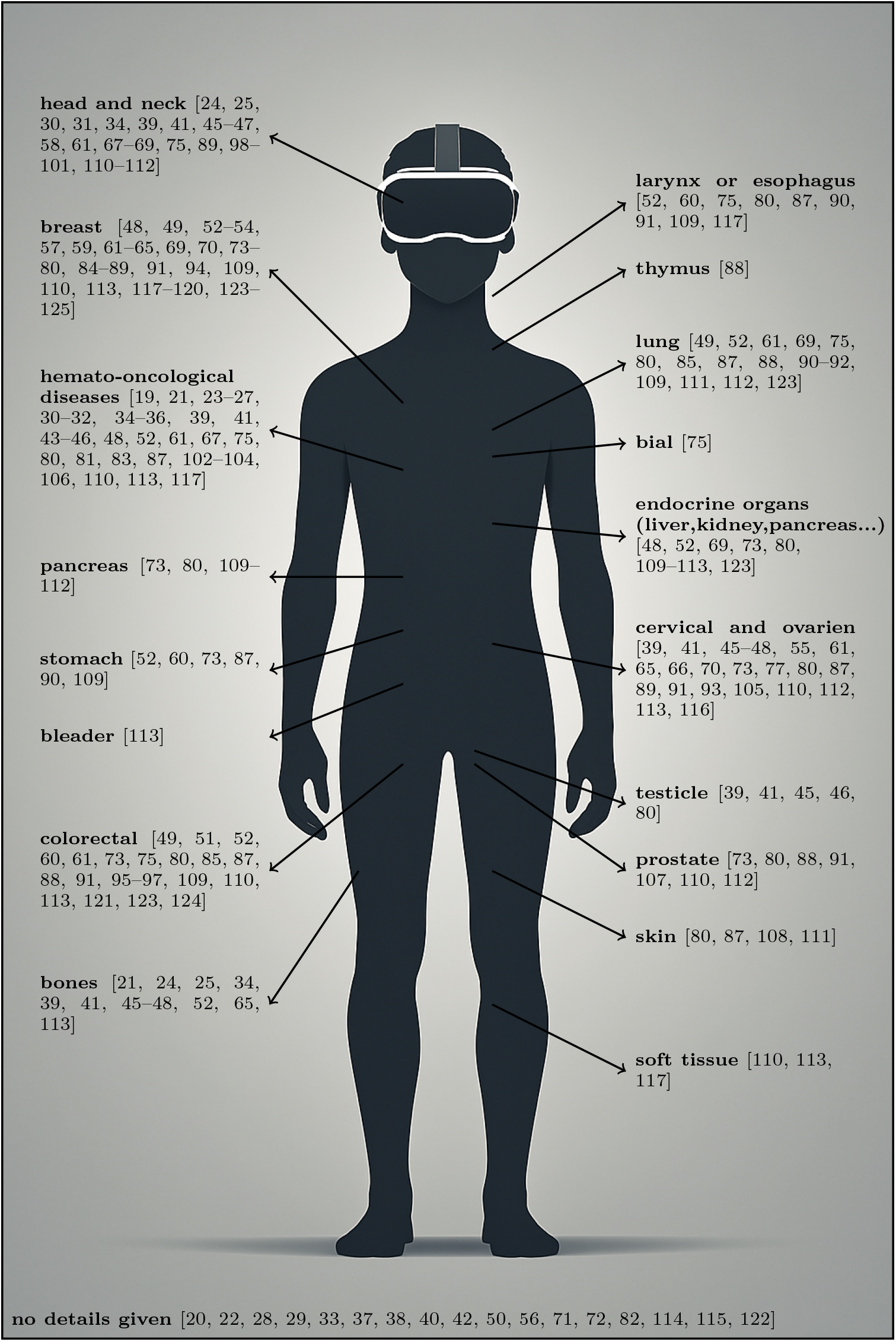
Distribution of the publications across various types of cancer.

In order to visualise the development of research over the years, the publications were also sorted by year of publication (Figure 4). Figure 4 shows a clear trend in the number of VR-related publications in the field of oncology over the last few years. The first report dates back to 1999. Until 2017, publications were rather sporadic, while from 2017 onward there was an almost exponential increase in publications.

**Figure 4:**
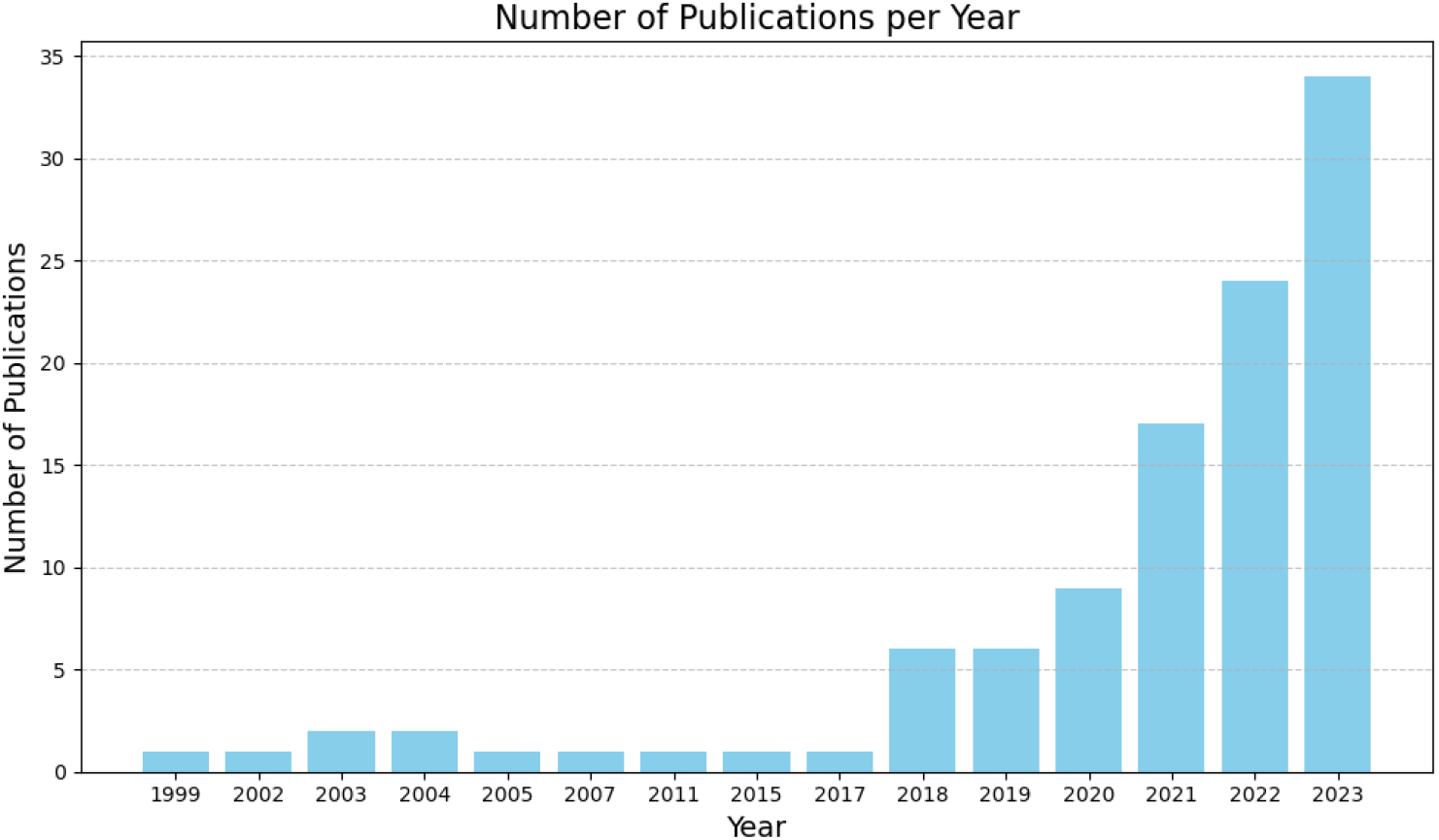
Distribution of publications sorted by year of publication.

If the publications are categorized according to the country in which they were conducted, it can be seen that around a quarter of the publications came from the United States, followed by China, Italy, France, Australia, and Canada. China, Italy, and France together account for around a quarter of all publications, with 11.3, 8.5, and 7.5%, respectively. Australia is also represented with 7.5%, Canada with 5.7, Turkey with 4.7 and Germany with 3.8%. All other countries account for less than 3%. The countries that accounted for less than 1% were grouped in the category ‘others’ for visualization.

To get an overview of the different hardware, a table was also created listing the different HMDs, the number of studies that used them, and the studies themselves (Table 3). Some studies used 2 devices, so they are listed more than once. Several devices of this brand were also grouped together under PICO and Sony HMDs, because detailed information about the generation of the device was only partially available, and to keep the table as small and clear as possible. HMDs that only appeared once have been grouped together in the ‘other’ category for the sake of clarity. In addition, 23 of the 107 publications did not provide information on the hardware used.

**Table 3:**
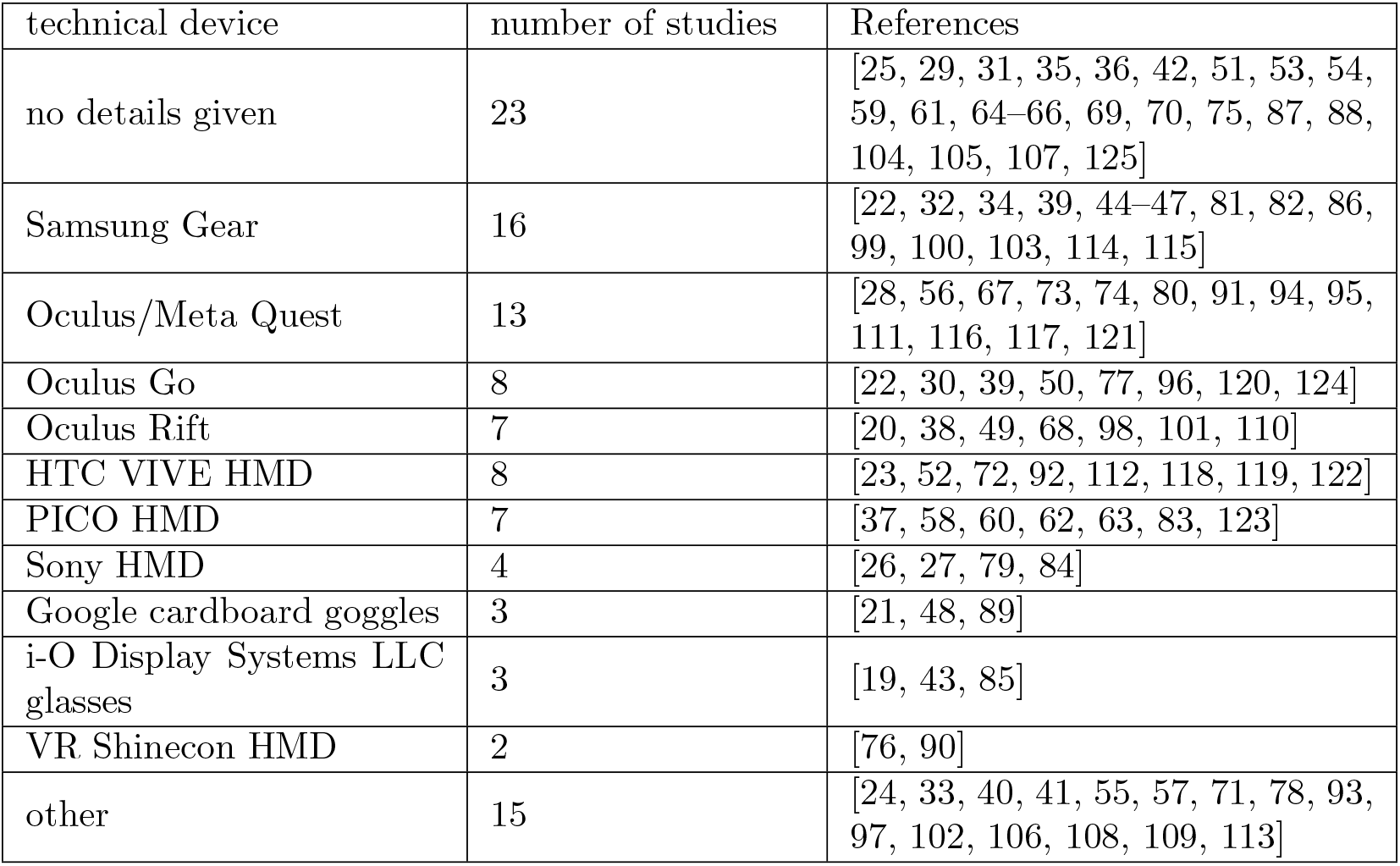
Number of reviews and listed reviews per VR device.

| technical device | number of studies | References |
| --- | --- | --- |
| no details given | 23 | [25, 29, 31, 35, 36, 42, 51, 53, 54, 59, 61, 64–66, 69, 70, 75, 87, 88, 104, 105, 107, 125] |
| Samsung Gear | 16 | [22, 32, 34, 39, 44–47, 81, 82, 86, 99, 100, 103, 114, 115] |
| Oculus/Meta Quest | 13 | [28, 56, 67, 73, 74, 80, 91, 94, 95, 111, 116, 117, 121] |
| Oculus Go | 8 | [22, 30, 39, 50, 77, 96, 120, 124] |
| Oculus Rift | 7 | [20, 38, 49, 68, 98, 101, 110] |
| HTC VIVE HMD | 8 | [23, 52, 72, 92, 112, 118, 119, 122] |
| PICO HMD | 7 | [37, 58, 60, 62, 63, 83, 123] |
| Sony HMD | 4 | [26, 27, 79, 84] |
| Google cardboard goggles | 3 | [21, 48, 89] |
| i-O Display Systems LLC glasses | 3 | [19, 43, 85] |
| VR Shinecon HMD | 2 | [76, 90] |
| other | 15 | [24, 33, 40, 41, 55, 57, 71, 78, 93, 97, 102, 106, 108, 109, 113] |

### 3.3 Research Questions

The overall aim of this systematic review is to analyse studies published up to the end of 2024 on the use of VR in the field of oncology. The main research questions for our study are as follows:

1. What are the current applications, developments, procedures, research areas and challenges in the use of VR in oncology for patient-centred care during therapy for all cancer types and stages? 2. What advantages does the use of VR technology together with conventional therapies in oncology offer for patient-centred care compared to conventional therapies, e.g., in terms of patient comfort and the reduction of side effects?

### 3.4 Outline and Organization of Reviews

The areas of application for VR in the field of oncology are diverse and the study groups, measured parameters, and hardware used vary broadly and are pretty heterogeneous. In order to summarise several studies under one rubric, the categories of application area were finally established, which were intended to represent groups that were as homogeneous as possible. 27% of the studies were conducted in pediatric oncology clinics, which is why a separate category was created for studies in children and adolescents. The group of adults was subdivided into smaller sub-categories for the sake of clarity. A sub-category based on age alone would have made the group too large and heterogeneous.

The time of VR application in the course of therapy and the type of therapy in detail (chemotherapy, radiotherapy, surgery, etc.), as well as the aim of the application were used for further subdivision. Figure 2 shows the taxonomy of subdivision into the specific categories. The subcategories are general cancer treatment (if no details on timing and treatment are given), during chemotherapy, before therapy, during surgical/invasive procedures, palliative treatment, rehabilitation after surgery, and detection of side effects. Table 2 shows the number of publications per subgroup and lists them. The individual chapters are listed below in order of the number of studies in the respective subcategory.

#### 3.4.1 Children

29 reports out of a total of 107 reports focused on VR in relation to children and adolescents. This represents the largest of the nine groups. Children and adolescents show a particularly high affinity for technology and are particularly easily distracted, and on the other hand are particularly vulnerable and susceptible to negative psycho-emotional effects from the disease and treatments. This makes them a rewarding target group for VR distraction experiments. It has already been proven that children undergoing cancer treatment in particular experience a high level of psychological stress [18], which is why there has been a lot of research in this area for the last decades. While distraction has been used previously in children receiving chemotherapy, Schneider and Workman used VR as an application for this purpose as early as 1999 [19]. The overall aim of this study was to reduce side effects such as anxiety, nausea, and vomiting during chemotherapy. For this purpose, VR distraction was used during the second course of chemotherapy, and patients’ sensations during the first three chemotherapy sessions before, shortly after and 48 hours after chemotherapy were recorded. Using a Virtual i-O brand headset and child-friendly videos on CD-ROM, patients showed a significant reduction in stress levels during chemotherapy with VR application compared to the two chemotherapies without application. However, the long-term effect of the VR application could not be ascertained. VR technology is developing rapidly, and current VR applications are much more immersive thanks to higher resolution displays, improved tracking systems, and advanced hand-tracking, giving users a stronger sense of a second reality in which they find themselves. In 2018, Ng et al. programmed an entire game for the Oculus Rift DK2 to distract children during chemotherapy with a virtual farm game [20]. The development of low-cost, disposable HMDs, such as the disposable Google Cardboard VR goggles, is also a great benefit for clinical use with oncology patients, as it minimizes the risk of infection, reduces costs, and allows users to use their own phones, bringing the point of care into the home situation. Wong et al. showed a significant reduction of anxiety and nausea after chemotherapy in a study with children age 6-12 undergoing their first chemotherapy using these goggles by measuring anxiety via the State Anxiety Scale for Children (CSAS-C) and acute nausea and vomiting during chemotherapy via the MASCC Antiemesis Tool. VR was applied several times over the first two days of chemotherapy, and parents, nurses, and their perspectives on VR interventions in pediatric oncology were included in the study. [21]. Erdős and Horváth only partially confirmed these results in their study with 10-18 year old children using a VR application during the first chemotherapy. Here, VR was only used once before the first treatment. Overall, the test subjects here also showed less anxiety and an overall better mood, but no significant changes in perceived nausea. [22]. The largest area of application for VR in pediatric oncology is the reduction of pain: Chiu et al. successfully used an HMD with interactive nature images to distract children with acute lymphoblastic leukemia from their pain during L-asparaginase chemotherapy (a painful injection into the muscle) [23]. However, the effect of VR on the perception of pain during minimally invasive procedures and punctures such as venipuncture and peripheral intravenous cannulation [24–28], port access [29–36], central venous catheter dressing [37] or lumbar punctures [43] was mainly investigated and showed overall significantly reduced pain perception in children and adolescent patients. Hoag et al. tested with a heterogeneous group of children age 8-25 receiving a mixture of the listed invasive procedures. The authors reported that some children wanted to be able to see their surroundings and the procedure and therefore felt anxiety during the VR application, indicating that the success of VR distraction during intravenous applications can vary greatly depending on a patient’s age and specific treatment [44]. These results are supported by Tennant et al. in a study on increasing psychological overall well-being in children receiving cancer therapy, including a verification stage to validate different VR software in comparison to a 2D video experience on a tablet [45, 46]. It has also been shown that VR therapies have a long-term effect on adolescents’ perception of pain when the technique is applied over a longer period of time and not just at the time of the direct pain stimulus [47]. The software used was exclusively nature videos (e.g., jungle, underwater, snow), cartoons, or roller coaster rides. Unfortunately, often no information was given about the hardware used. Releases around the turn of the millennium use Virtual i-O brand headsets [19, 43]. More recent publications mainly use Samsung HMDs [32, 34, 44–47] and the Oculus Quest [28, 30]. Sony HMZ T-2 glasses [26, 27], Pico Goblin glasses [37] and the Piranha™ VR system [33] were also used. Another area of application was the preparation for radiotherapy sessions. Radiotherapy is a treatment method that does not involve visible procedures like surgery or medication intake, which makes it more abstract and difficult for children and adolescents to comprehend. Patients have to lie alone and as motionless as possible in a device for a long period of time, which is particularly difficult for children and causes great anxiety. Galvez et al. designed a 360 degree video VR-tour of a radiotherapy room and provided children with an immersive experience through the Oculus Rift Development Kit 2 headset prior to their own therapy. The patients reported that this tour made them feel calmer before their first radiotherapy treatment [38]. Tennant et al. were able to confirm these results with a similar application on a Samsung headset [39]. Schenck et al. developed a game to teach children radiotherapy in a playful way using Google VR SDK glasses in conjunction with a cell phone and were also able to increase patients’ understanding and reduce their anxiety on a significant level [40]. In addition to the classic VR applications in the form of HMDs, there is a study by Li et al. in which a VR was generated in the form of 3D projections on walls and floors by a so called PlayMotion!^®^ system. The children were able to interact with the immersive underwater world using the shadows of their arms. This study also showed a significant reduction in depressive symptoms in the children [41]. Usó, Sandnes, and Medola went one step further and had the children draw monsters, aliens and underwater creatures in a specially designed VR environment on the HMD VR Box and created real-life toys from these drawings. The children thoroughly enjoyed the project and also showed a significantly reduced pain response and less fear [42].

#### 3.4.2 During cancer treatment in general

While section 3.4.1. comprises studies on pediatric patients, the following sections 3.4.2-3.4.8 focus on adult patients. These studies on adult patients are spread across several areas of application in the broad field of oncology. Generally, accompanying therapy was the next major area of VR applications for cancer patients, accounting for 23 of the 107 studies.. VR was mainly used to reduce pain [48–54] or psychological symptoms such as anxiety and depression [53, 55–65] in cancer patients. In some studies, VR applications were used in combination with conventional therapies for reducing side effects, such as in combination with morphine [53] or tramadol [54] to reduce the dose of painkillers with the same result if possible, or to teach pain self-efficacy to patients using progressive muscle relaxation and guided pain visualization techniques, to make the administration of painkillers completely obsolete in the best case [49]. Sometimes VR was also used together with psychological applications such as cognitive behavioral therapy [66] or mindfulness-based stress reduction [65], as a virtual meeting room of a self-help group with other patients [67], or simply as visual stimulation during psychotherapeutic application [59]. Some studies also went beyond psychological applications and even used VR to treat other problems such as cognitive impairment [68, 69] and hot flashes by delivering a nature based snow experience called Frosty [70]. The most commonly used hardware was the Pico Goblin VR headset [58, 60, 62, 63] and the most common software was nature-based relaxation recordings [50, 51, 55, 57, 59, 64, 65]. Song et al. even tested nature recordings in comparison to other urban environments in their study of 63 patients with oesophageal and other gastrointestinal cancer and an average age of around 60 years, with nature recordings producing the best results [60]. Chin et al. confirmed these results in their study with 38 women with metastatic breast cancer, detecting significantly reduced fatigue and depression and increased quality of life. They could also show a correlation between particularly good results in reducing depression and a stronger sense of connection to nature among the patients [62]. Regarding the hardware, HMDs for mobile phones have been particularly widely used [53, 70]. Among other things, Google Carboard[48] and the VR headset from iHarbot^©^ [57] were used. There were also studies using the Oculus Go [50], the Oculus Rift [68] and the Meta Quest 2 [56]. VR was used together with heart rate monitors in two studies. Rolbiecki et al. used the Oculus Go together with Brainlink, a portable electroencephalography, in their study with 15 patients with cancer related pain to visualize the patient’s pulse in the form of a dragonfly when pain is detected in VR and to enable the patient to reduce their pulse independently and in a targeted manner, with the result that both pain was significantly reduced and the patients’ perceived depression and anxiety decreased [50]. Matsangidou et al. had a similar approach and used the Google Cardboard HMD together with a smartphone and a Garmin smartwatch to track the patient’s pulse in their study containing 23 cancer patients and 26 medical and paramedical personnel. This should enable the patient to independently start a VR application if their pulse rate increases after perceiving pain or anxiety, showing that it can be efficacious and safe for cancer patients for improving symptoms related to pain, fatigue, shortness of breathing, depression, anxiety, and general well-being [48]. In addition, the BOBO VR Z4^®^ HMD [55] and the HTC Vive stereoscopic headset [52] were used in two further publications.

#### 3.4.3 During Chemotherapy

The third largest area of application was during chemotherapy, with 15 of the 107 studies. The focus here was almost exclusively on reducing psychological side effects such as depression, anxiety, stress and fatigue. In a few studies, the patients’ perception of pain was also assessed in order to define possible applications here [71–73]. As in the previous chapter, VR apps based on nature images were used in most of the studies [71–80, 83, 85]. In addition, some studies also used apps with a background in culture, such as museum visits or visits to historical sites [77, 79, 85]. A VR game was also specially designed for a study in which 26 middle-aged cancer patients at the Department of Hematology, Blood Cancer, and Bone Marrow Transplantation were able to shoot at cancer cells in the style of a first-person shooter game with the help of an avatar in an abstract environment [81]. Burrai et al. focused not only on chemotherapy patients, but also on patients receiving infusions of other anti-neoplastic therapy, and compared VR with narrative medicine to reduce anxiety and fatigue during administration. 74 middle aged patients were split into three equal sized groups for this study to compare both techniques with the standard care. Narrative medicine is an approach in healthcare that emphasizes understanding patients’ personal stories and experiences as part of their treatment, aiming to improve empathy, communication, and patient-centered care. It encourages healthcare providers to listen actively and incorporate patients’ narratives into their medical care to foster a deeper therapeutic connection. Both techniques showed a significant improvement in the measured anxiety levels compared to the control group, which received standard treatment, with the VR group showing better results in comparison. Fatigue was only significantly reduced in the VR group [80]. In most studies, the effects of the VR application were measured using questionnaires and established scores derived from them. Ashley Verzwyvelt et al. also measured the saliva cortisol level of their 36 first-time Chemotherapy patients to obtain information about the stress level of the patients [73].

Interestingly, in seven out of the 15 studies, no cybersickness or at least no significant increase in nausea was measured [72, 74, 75, 79, 80, 84, 85]. However, many studies did not provide any information about cybersickness symptoms at all [71, 73, 76, 77, 81, 82]. Two studies also paid special attention to this risk and deliberately chose short VR applications with little movement [76, 77]. Only Chirico et al. reported a cybersickness rate of less than 20 percent in their 94-person study of breast cancer patients and Zhang et al. reported one patient with acute symptoms and 5 with mild dizziness in their 63-person study of leukaemia patients [78, 83]. In two studies, the Samsung Gear VR HMD together with a smartphone were used [81, 82] and even more studies used the Oculus Quest 2 [73, 74, 80]. Other hardware choices included the VR One HMD by Zeiss [71], Zore G04BS VR Shinecon VR HMD with smartphone [76], Oculus Go [77], HTC Vive^®^ VR HMD [72], Vuzix Wrap 1200 VR HMD [78], the i-Glasses^®^ SVGA HMD [85], the Pico Neo 3 Pro - 6 DoF [83] and the Sony PC Glasstron PLM-S700 headset [79]. In summary, none of the studies showed a significant negative effect of VR on the patients. Some showed a positive effect, especially in terms of general distraction and relaxation for patients [71, 72, 75– 77, 83, 84]. Still many studies did not show statistical significant improvement in the measured parameters [73, 79, 81, 85] or without significant improvement or just minimal improvement compared to standard applications for distracting patients such as music [74, 78].

#### 3.4.4 Before therapy

The fourth largest area of application was the use before the start of therapy to prepare patients for the various treatments and reduce their anxiety. This area of application accounted for 13 of the 107 studies. The reports focus on preparation for chemotherapy [86, 87], preparation for radiotherapy [88–94] and preparation for surgery [95–98]. In both studies on the use of VR before the first chemotherapy, instructional videos were used to show and explain the chemotherapy procedure in advance. The success of the application was measured by the use of established scores based on surveys, for example to classify anxiety. Torres García et al. conducted a study on 113 breast cancer patients and evaluated emotional state, anxiety and depression levels using the Hospital Anxiety and Depression Scale (HAD), coping levels using the Mental Adjustment to Cancer scale (MINI-MAC), and emotional distress using the Differential Emotions scale (DME). A significant improvement was demonstrated in all areas compared to the control group [86]. Birkhoff also used a VR information video before the first chemotherapy and tested their experimental setup on 35 cancer patients receiving their first chemotherapy without a control group. They measured heart rate, blood pressure, and anxiety via the State-Trait Anxiety Inventory (STAI) and were able to determine a significant reduction in every item compared to the measured baseline before the application [87]. In the four studies on the use of VR before surgery, three reports worked with colorectal cancer patients [95–97] and one with brain tumor patients [98]. While Turrado et al. and Shepherd, Trinder, and Theophilus relied on instructional videos with 3D simulations of the surgical procedure in their studies [95, 97], Schrempf et al. used relaxation apps to increase the well-being of patients before and shortly after colorectal cancer tumour resection through several applications [96]. In their study with 33 brain tumor patients prior to surgery Perin et al. also used instructional videos in VR for 3D visualisation of the procedure in order to achieve a better understanding of the procedure and a reduction in anxiety. For this purpose, they used DICOM files of patient CT scans of the affected regions. However, compared to the control group, only the improved objective understanding could be demonstrated and no significant reduction in anxiety [98]. In the seven studies on application before radiotherapy [88–94], only instructional videos were used. While most of the studies focused on well-being and the reduction of anxiety in patients, Zhao et al. also evaluated the effects of the application on the occurrence of set-up errors during treatment and found an improvement both here and in patient well-being in their study on 120 cancer patients with thoracic- or abdominal cancer before their first radiotherapy [90]. Johnson et al. have also validated their VR application in patients who have already received radiotherapy to get feedback on improvements compared to the previous approach. The aim was to improve patient education for radiotherapy applications in the long term. [93]. Not all publications gave details regarding the hardware used, but among others the used HMDs were the Samsung Gear device [86], the Oculus Quest [91, 94, 95], Oculus Go [96] and Oculus Rift [98], Google cardboard VR viewer [89] or Bluebee™ Genuine VR 3D Glasses [97] with smartphone, a HTC VIVE system [92]and the View-Master VR headset [93].

#### 3.4.5 During a surgical/invasive procedure

Eleven studies investigated the use of VR during surgical procedures. A large proportion of these are studies which used VR for brain mapping during awake brain surgery [99–101]. Brain mapping during brain surgery is a technique used to identify and preserve critical brain areas responsible for functions like movement, speech, and sensation, helping to minimize the risk of neurological damage while safely removing tumors or abnormal tissue. Five other studies evaluated the pain and anxiety reduction of patients during applications such as bone marrow biopsy [102, 103] or hematopoietic stem cell transplantation [104], uterovaginal brachytherapy applicators’ removal [105] and lumbar puncture [106]. In two studies on the use of VR for brain mapping, a 3D VR version of the already established picture-naming task, DO 80 [126], the most established exercise for language mapping was used [99, 100]. Cybersickness was documented and no significant increase was found. VR was well accepted by the patients. However, cases of intraoperative focal seizures were documented in both studies, but these were not necessarily related to VR. Casanova et al. used a VR application where patients should recognize the person looking at them out of a group of faces and define their emotion based on their facial expression. The goal was to check if the patients have difficulties exploring the space, locating the face of the avatar looking at them, or fail to recognize the facial emotion. The study was performed on 15 adult patients, hospitalized for a brain tumor near language and/or motor areas [99]. Mazerand et al., however, used a VR version of the Estermann test to assess the patients’ visual impairment during surgery in their study on ten adult patients presented with visual field defects that were undergoing awake brain surgery. The VR system detected 90% of the visual field defects found by automated perimetry, demonstrating a high level of accuracy in identifying visual impairments. One patient who underwent awake craniotomy with the VR system had no permanent postoperative visual field defect, confirming the potential utility of the system in preserving visual function during surgery [101]. Only interactive nature images were used as a distraction during lumbar puncture, uterovaginal brachytherapy applicators’ removal, bone marrow biopsy and hematopoietic stem cell transplantation [102– 106]. Wong et al. on the other hand performed a study in which VR hypnosis is used for pain and anxiety reduction during prostate biopsy or gold seed insertion procedures on 23 patients [107]. Menekli, Yaprak, and Doğan used distraction by VR nature images to reduce pain and anxiety and influence vital signs of cancer patients during venous port implantation. With 139 patients, their study represents the largest number of participants in this section and is also the only one which used a control group. They were able to show that the use of VR reduced the patients’ pain, anxiety, systolic blood pressure, diastolic blood pressure, heart rate and respiratory rate and increased oxygen saturation compared to the control group [109]. Higgins et al. also used VR nature recordings after Mohs surgeries for 116 patients with skin cancer to reduce patient anxiety with the result that VR experiences during the Mohs surgical day significantly improved anxiety and patient satisfaction [108]. Not all authors gave details regarding the hardware used, but among others the used HMDs were the Turkcell T-VR glasses [109], HTC Vive headset [99], Samsung Gear HMD with Samsung S7 smartphone [100], a Vive VR headset and Google Daydream headset [108], a Sony PlayStation VR2 headset [106], zVision X4 virtual reality goggles [102] and two different HMDs from Oculus [101].

#### 3.4.6 Palliative care

Eight publications dealt with the use of VR in the field of palliative care for cancer patients. The aim of the applications was exclusively the reduction of symptoms such as pain, depression, and anxiety. The majority of the studies therefore used relaxing nature apps such as Nature Treks VR on the Oculus Rift [110] or the Oculus Quest 2 [111], or the HTC VIVE HMD together with the Google Earth VR software [112], or the Mirage Solo VR HMD in combination with extra designed interactive and non interactive nature videos and games [113], or Samsung Gear HMD [114] and PICO G2 4K HMD [115] together with various nature and animal images. Some studies also focused on a personal connection between the patients and the images shown and either used nature images of the region [116] or asked the relatives to take images of themselves and the patient’s familiar surroundings and linked these to telephone conversations with the relatives during the VR application [117].These two studies also used the Oculus Quest 2 for their applications. In most studies, the usual scaling methods for measuring perceived anxiety, depression or pain were used and generally promising results were achieved. The use of HMDs in bedridden patients showed a particularly great advantage in this area.

#### 3.4.7 Rehabilitation after a procedure

Five of the 107 studies dealt with the use of VR for rehabilitation after therapy. Of these, four used VR for rehabilitation after surgery [118–121]. The last of the five publications dealt with the psychological rehabilitation of cancer patients [122]. Persson et al. designed a virtual smash room in which patients could vent their anger and resentment after therapy and tested it on 101 patients in a cancer rehabilitation center [122]. The study participants enjoyed the VR application, with only slightly more than 20% experiencing mild symptoms of dizziness, although these could not be directly linked to the VR application. Zhou et al. validated their VR rehabilitation system directly in two studies in 20 and 15 breast cancer patients post surgery [118, 119] with no significantly increased cybersickness symptoms detected. Buche et al. used a test period of 10 months to ensure the long-term tolerability of VR rehabilitation apps on 52 breast cancer patients after surgery, resulting in a cybersickness rate under 10% [120]. Zhou et al. designed an app for their own use that transfers simple physical movement into VR and used the HMD from HTC Vive [118, 119]. Schrempf et al. used the Holofit app from Holodia with the Oculus Quest 2 and also focused on physiological regeneration [121]. Buche et al., on the other hand, used the Nature Treks app with the Oculus Go and music therapy with a focus on anxiety reduction and mood enhancement [120]. In the self-designed application by Persson et al., the focus was also placed on mental rehabilitation and the HTC Vive VR system was used [122]. Overall, all studies showed promising results, albeit still in the experimental phase. Although great emphasis was placed on the identification of cybersickness symptoms and there were some reports of such symptoms, none of the five studies found an increase in such symptoms with statistical significance.

#### 3.4.8 Detection of side effects

In addition to the generally known side effects, such as nausea and hair loss, cancer therapies often have neurological and psychological side effects that are less prominent, such as a reduction in memory performance or balance disorders. That is why three publications focused on the measurement of side effects. Zeng et al. looked at the assessment of cognitive abilities of cancer patients in their study. They used a self-designed app and the ICO neo-3 portable VR HMD to assess verbal learning and memory, verbal fluency, information processing speed and executive function in their study on 165 cancer patients after primary cancer therapy. Participants’ performance on the VR cognition assessment showed a moderate to strong positive correlation with the results of their paper-and-pencil neurocognitive tests. The symptoms of cybersickness were negligible [123]. In their study,Teng et al. focused on the risks of chemotherapy-induced peripheral neuropathy (CIPN) on 34 breast and colorectal cancer patients and used a specially developed app and the Oculus Go VR HMD in combination with a Wii balance board to assess patients’ balance and to support and identify patients at risk of falling. The application showed very good results, comparable with existing methods [124]. Duivon et al. looked at the impairments in prospective memory (PM) and reduced sleep quality of 58 breast cancer patients under 70 years of age and at least one year post-menopause after hormone therapy. The sessions therefore took place at different times, including at night, and included memory exercises in a purpose-designed VR app. Their data suggest that the sleep-dependent mechanisms of prospective memory consolidation are not altered in early-stage breast cancer patients who are not treated with chemotherapy. At the same time, however, no negative side effects of VR use were reported [125].

## 4 Meta-Analysis

### 4.1 Extracting and Preparing Studies

In order to obtain a more structured overview of the results of the studies, a meta-analysis was carried out. For this purpose, all 107 papers were searched for the most frequently collected quantitative parameters. A minimum of ten studies was used as a cut-off. This resulted in two measured parameters usable for further analysis: STAI (State-Trait Anxiety Inventory) and VAS (visual analogue scale). First, the risk of bias (RoB) was assessed for all included studies using the current version of the Cochrane Risk of Bias 2 (RoB2) tool[127]. This tool evaluates potential sources of bias across five domains: (1) randomisation process, (2) deviations from the intended interventions, (3) missing outcome data, (4) measurement of the outcome, and (5) selection of the reported result. Each domain, as well as the overall risk of bias, was classified as low risk, some concerns, or high risk. Afterwards a random-effects meta-analysis was conducted. For this purpose, missing standard deviations were approximated according to the publication by[128]. 14 publications were therefore suitable for the final meta-analysis. Figure 6 shows the workflow for selecting and processing the studies.

**Figure 5:**
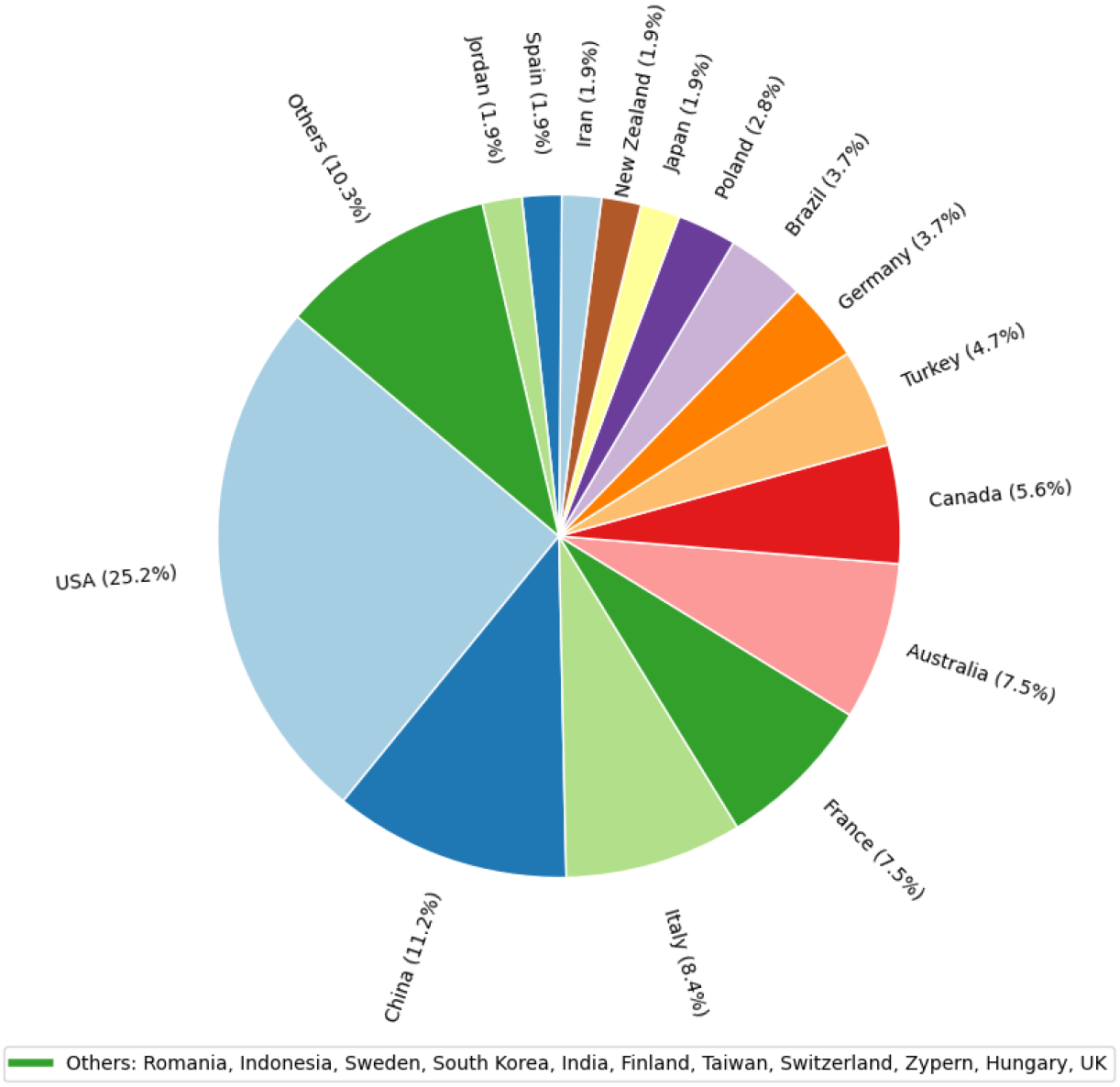
Distribution of publications sorted by country of conduction.

**Figure 6:**
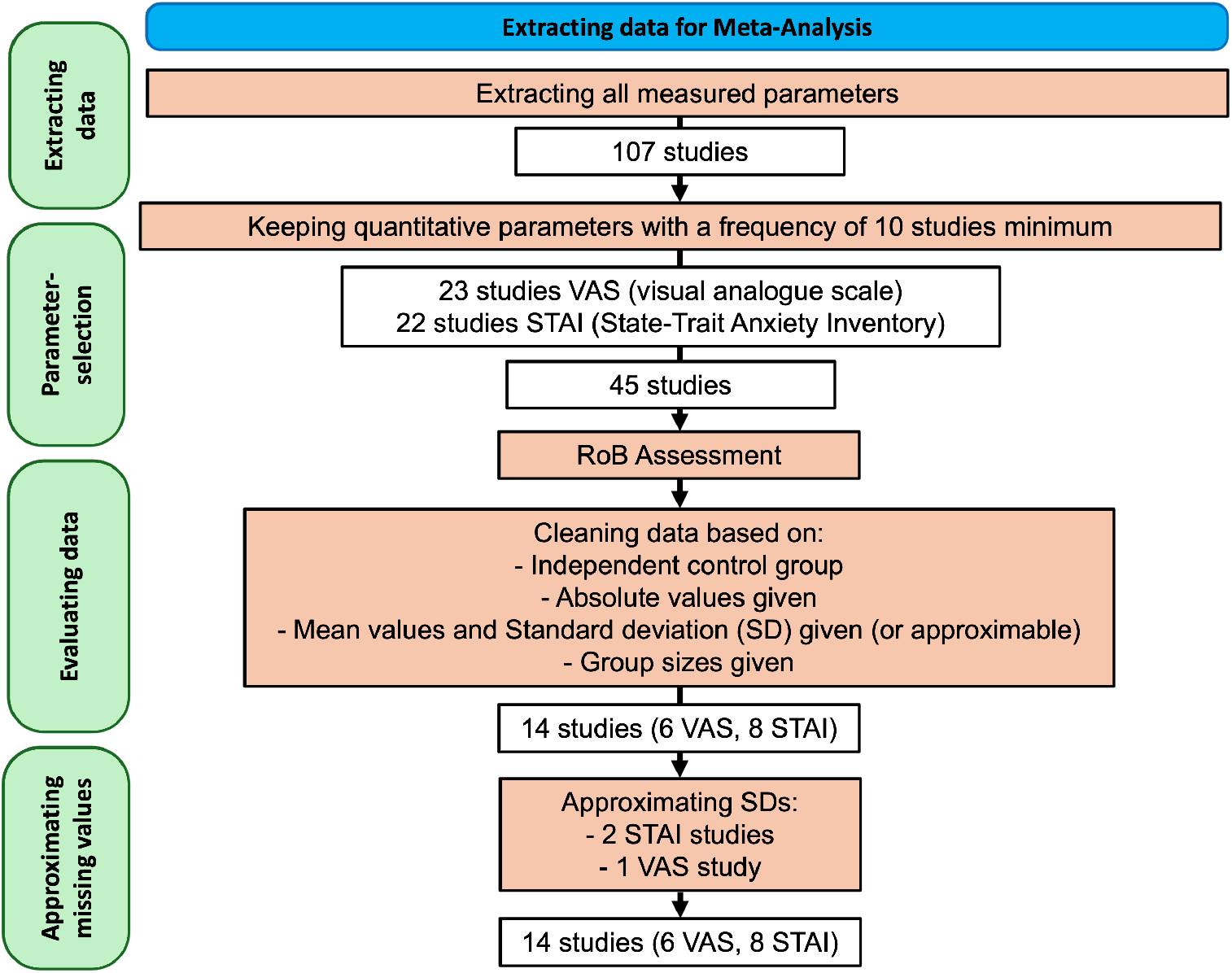
Search strategy used in this meta-analysis to extract reliable data. After removing reports with missing or non-absolute data and approximating missing standard deviations (SD) a total of 14 reports were used in the final analysis, splitted into two groups of six and eight reports.

### 4.2 Results Meta-Analysis

#### 4.2.1 risk of bias (RoB) Assessment

The risk of bias assessment was performed for all studies using parameters that were measured in a minimum of ten studies. Resulting in 2 figures (7 and 8) showing the distribution of risk across the five domains for the two parameters most frequently reported across studies: the State-Trait Anxiety Inventory (STAI) and the Visual Analogue Scale (VAS). STAI was measured in 22 studies and VAS in 23. For both outcomes, the overall risk of bias was rated as high in the majority of studies.

**Figure 7:**
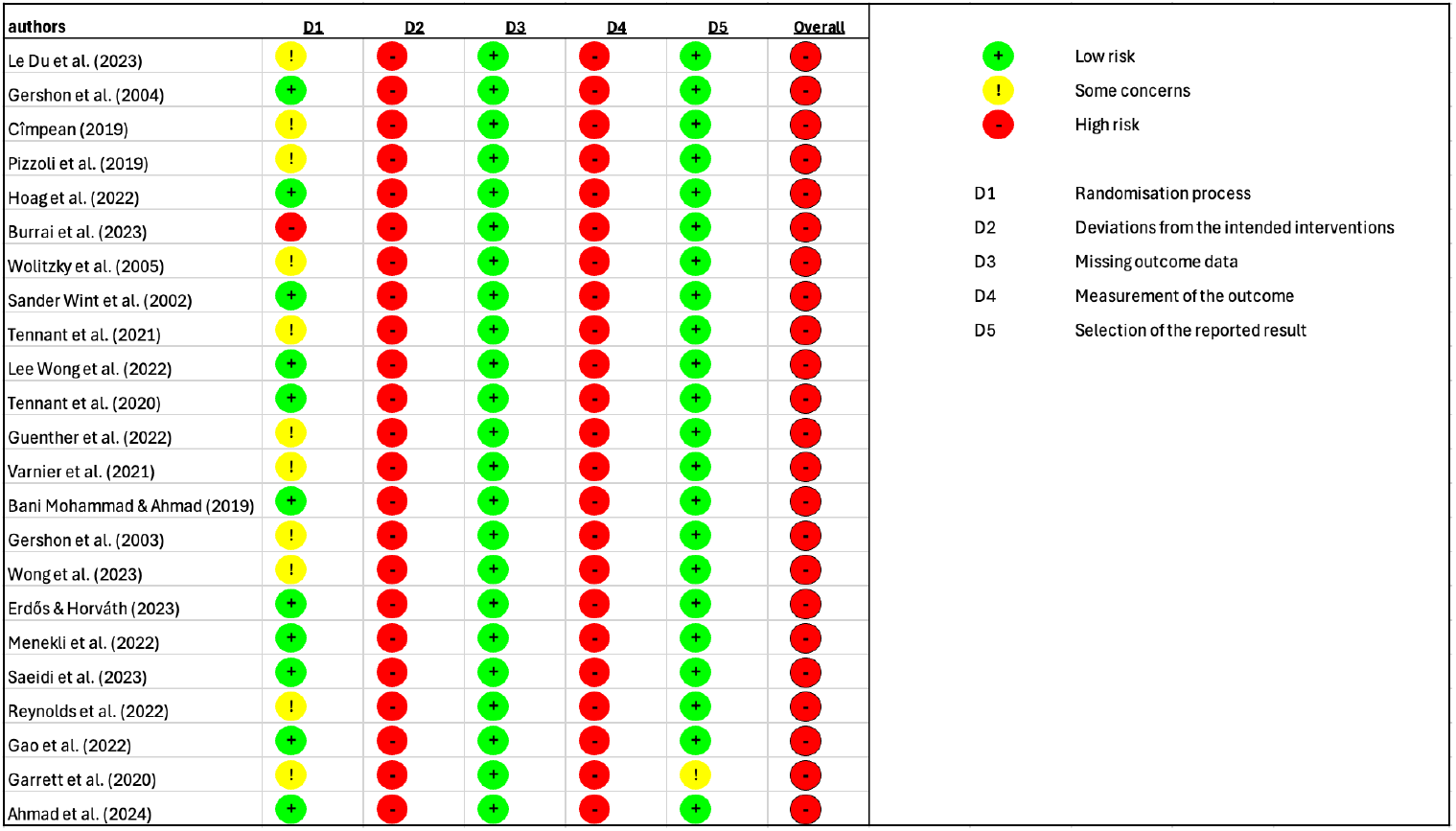
Risk of bias assessment for all studies using the visual analoguo scale (VAS) to measure the effects of VR usage. Tool evaluates potential sources of bias across five domains: (1) randomisation process, (2) deviations from the intended interventions, (3) missing outcome data, (4) measurement of the outcome, and (5) selection of the reported result and the overall risk of bias.

**Figure 8:**
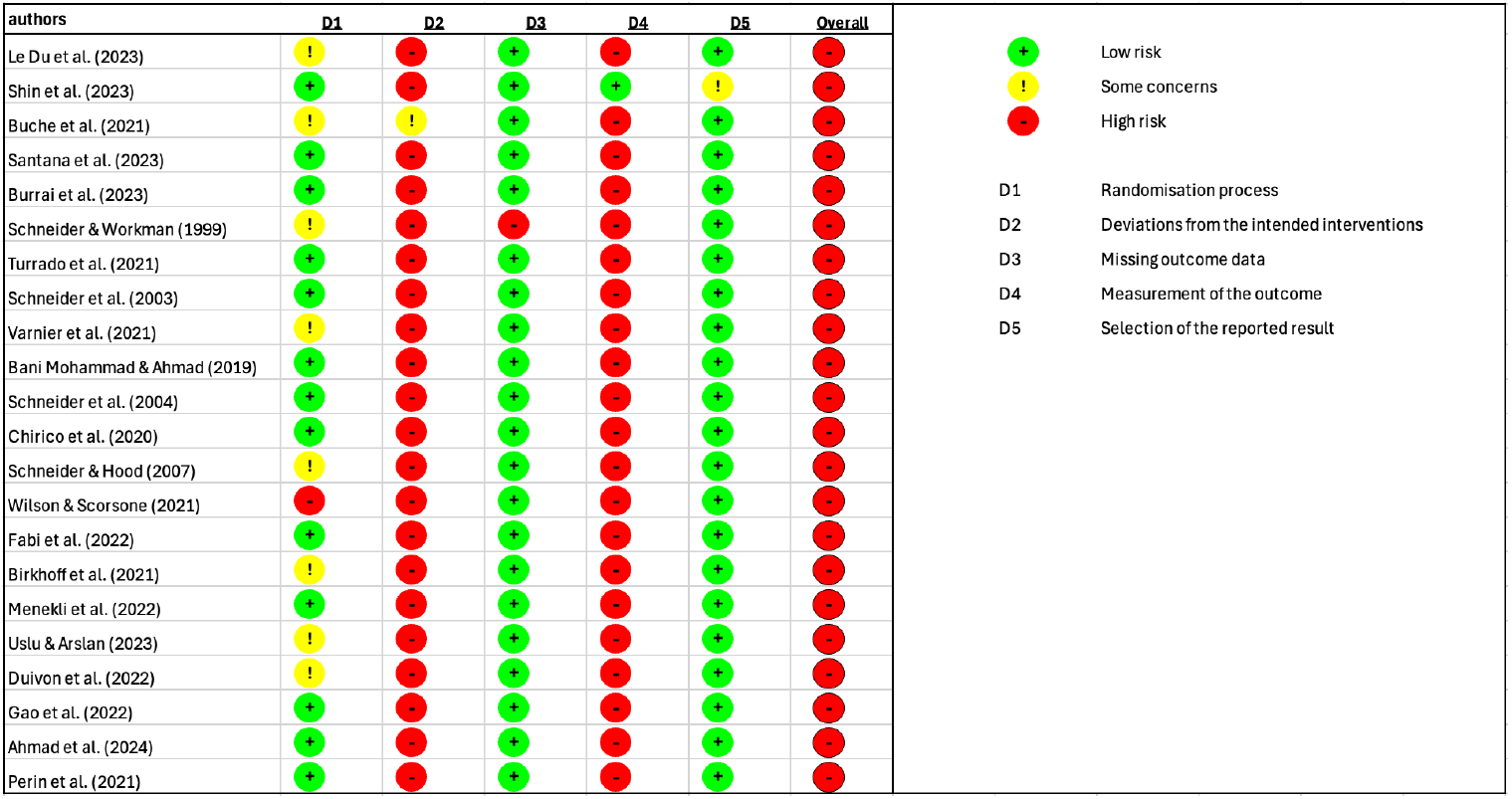
Risk of bias assessment for all studies using the State-Trait Anxiety Inventory (STAI) to measure the effects of VR usage. Tool evaluates potential sources of bias across five domains: (1) randomisation process, (2) deviations from the intended interventions, (3) missing outcome data, (4) measurement of the outcome, and (5) selection of the reported result and the overall risk of bias.

Most studies were rated as low risk in the domains of randomisation (D1), missing outcome data (D3) and selection of the reported result (D5) However, the domains of deviations from the intended interventions (D2) and measurement of the outcome (D4) showed frequent concerns, with many studies classified as high risk in these categories. No studies at all were rated as “low risk” across all domains. The visual presentation of the RoB plots highlights a similar pattern for both STAI and VAS: consistently high or unclear risk in multiple domains, resulting in a high overall risk rating for most studies.

#### 4.2.2 random-effects meta-analysis

A random-effects meta-analysis was conducted to evaluate the effect of the intervention on post-intervention STAI scores across eight studies, resulting in a funnel plot (figure: 9) and a forest plot (figure: 10) The pooled standardized mean difference (Hedges’ g) was –0.92 with a 95% confidence interval of [–1.89, 0.06], indicating a moderate-to-large effect in favor of the intervention. However, the confidence interval includes zero, suggesting that the overall effect did not reach conventional levels of statistical significance.

**Figure 9:**
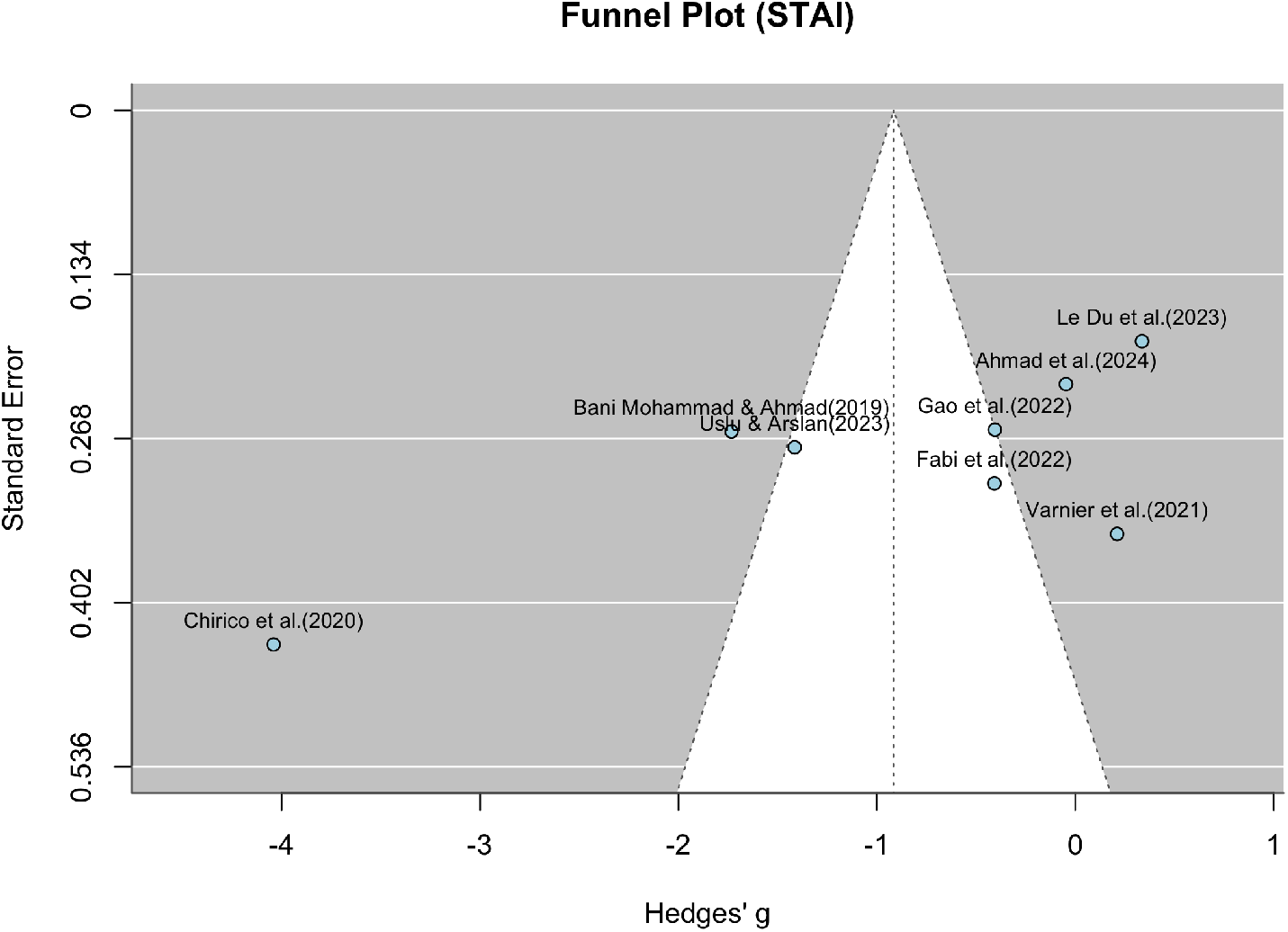
Funnel plot for STAI values. Shows the effect size of the individual studies in relation to their precision (study participants) for all eight studies that were suitable for the meta-analysis of STAI values.

**Figure 10:**
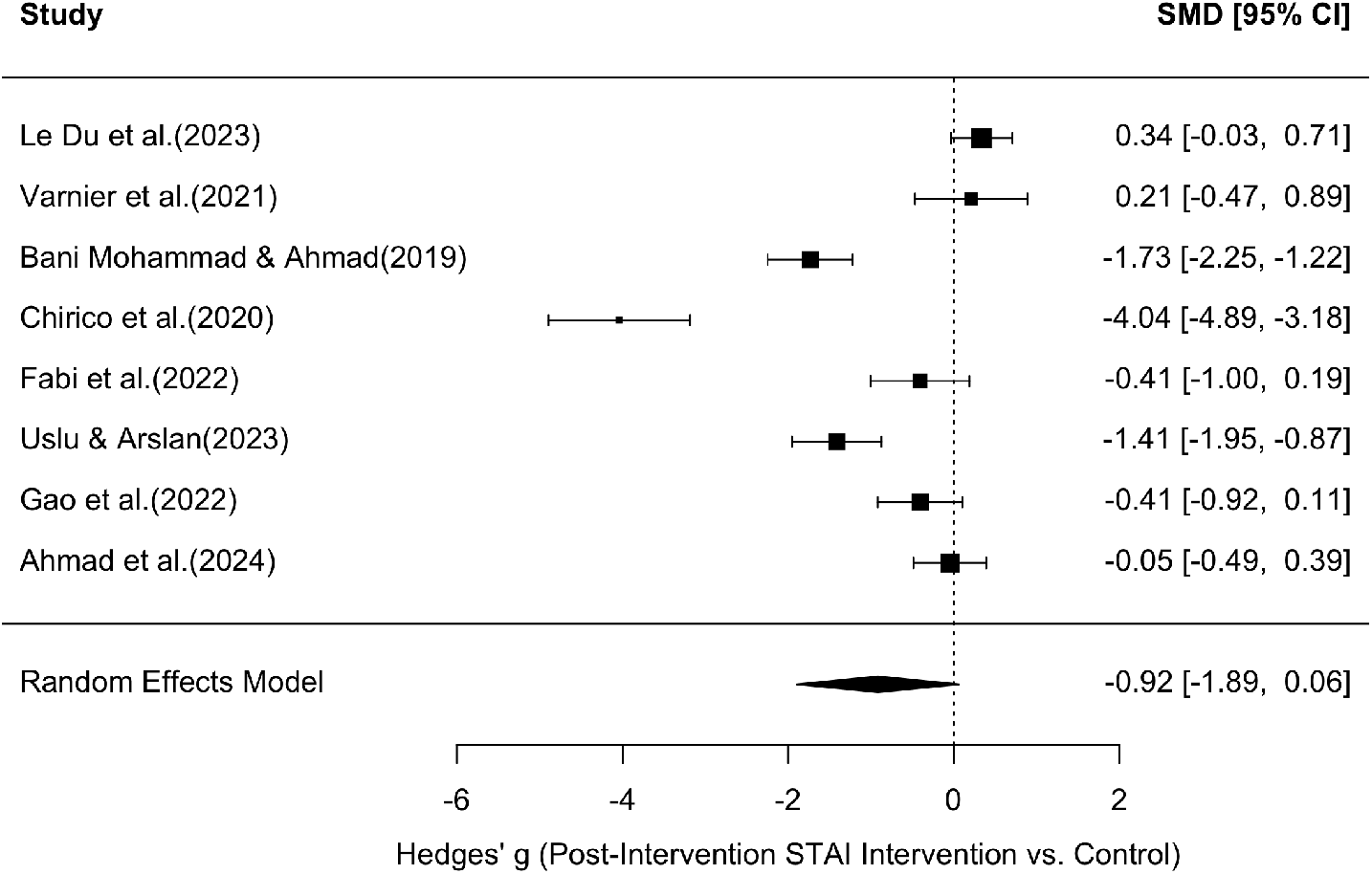
Forest plot for STAI values. Shows the results of the individual eight studies and the overall result of the meta-analysis for STAI values (random effect model), as well as the respective variance.

The forest plot (figure: 10) illustrates substantial variability in effect sizes across studies. Some studies, such as Chirico et al. (g = –4.04, 95% CI: –4.89 to –3.18), Mohammad and Ahmad, and Uslu and Arslan, reported strong negative effects (favoring the intervention), while others, such as Ahmad et al. and Fabi et al., reported small or near-null effects. The funnel plot (figure: 9) was visually inspected for asymmetry to assess potential publication bias. While the distribution of studies is roughly symmetrical around the pooled effect size, there is some suggestion of asymmetry, particularly due to one extreme outlier (Chirico et al.). This study showed a significantly larger effect size than all others with a simultaneously above-average standard error, which could indicate a statistical outlier. A random-effects meta-analysis was also conducted in the same way to assess the effect of the intervention on post-intervention VAS scores in six studies, resulting in a funnel plot (figure: 11) and a forest plot (figure: 12). The pooled standardized mean difference (Hedges’ g) was -1.02 with a 95% confidence interval of [-2.09, 0.05], which also indicates a moderate to large effect in favor of the intervention. However, the confidence interval also includes the value zero, which indicates that the overall effect did not reach the conventional level of statistical significance. The forest diagram (figure: 12) illustrates the considerable variability of the effect sizes between the studies. Some studies, such as Mohammad and Ahmad (g = -2.34, 95% CI: -2.91 to -1.77) and Menekli, Yaprak, and Doğan (g = -3.05, 95% CI: -3.53 to -2.56), reported strong negative effects (in favor of the intervention), while the remaining studies reported small or almost no effects(Ahmad et al., Cimpean, Le Du et al., Varnier et al.).

**Figure 11:**
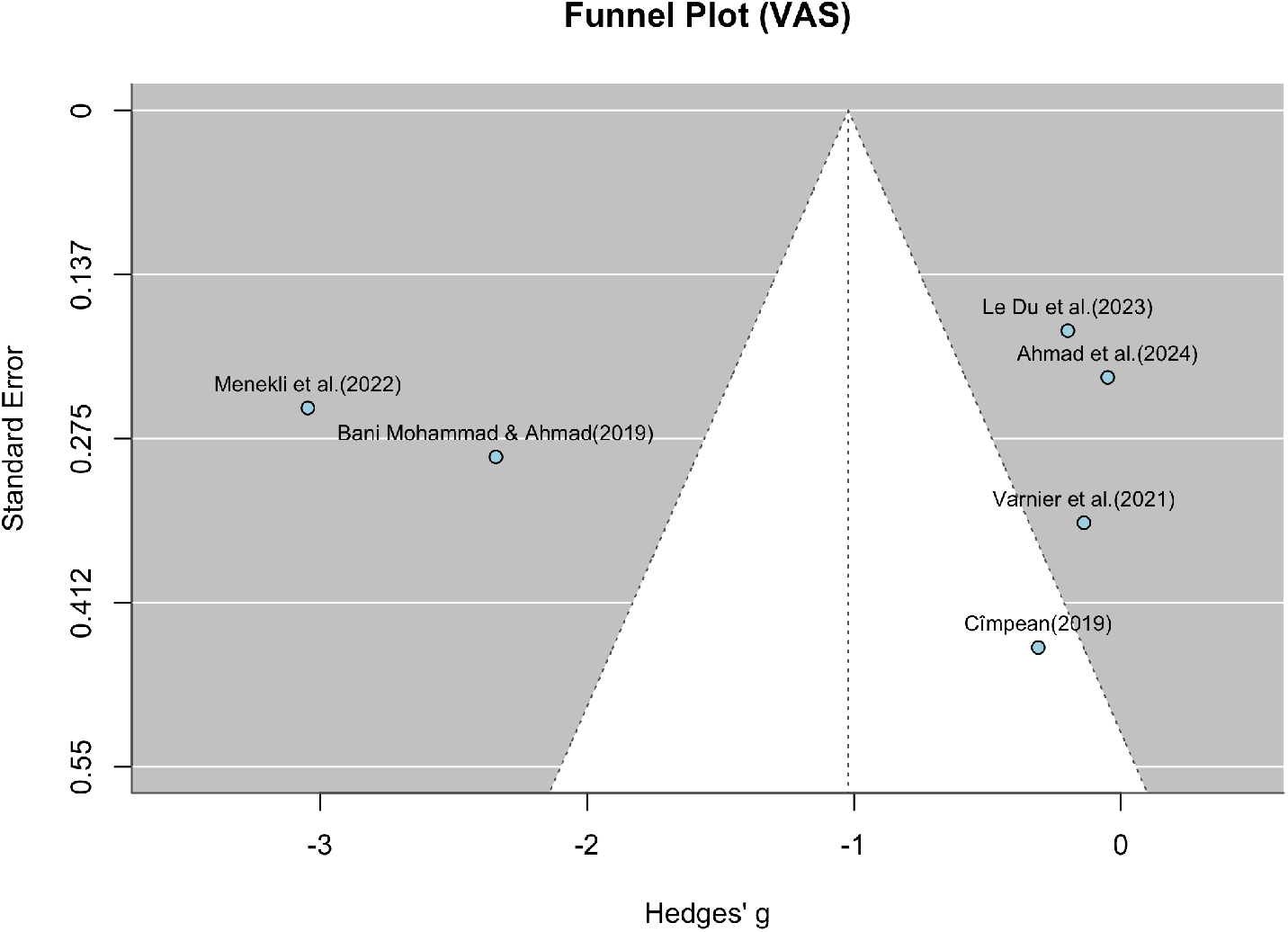
Funnel plot for VAS values. Shows the effect size of the individual studies in relation to their precision (study participants) for all six studies that were suitable for the meta-analysis of VAS values.

**Figure 12:**
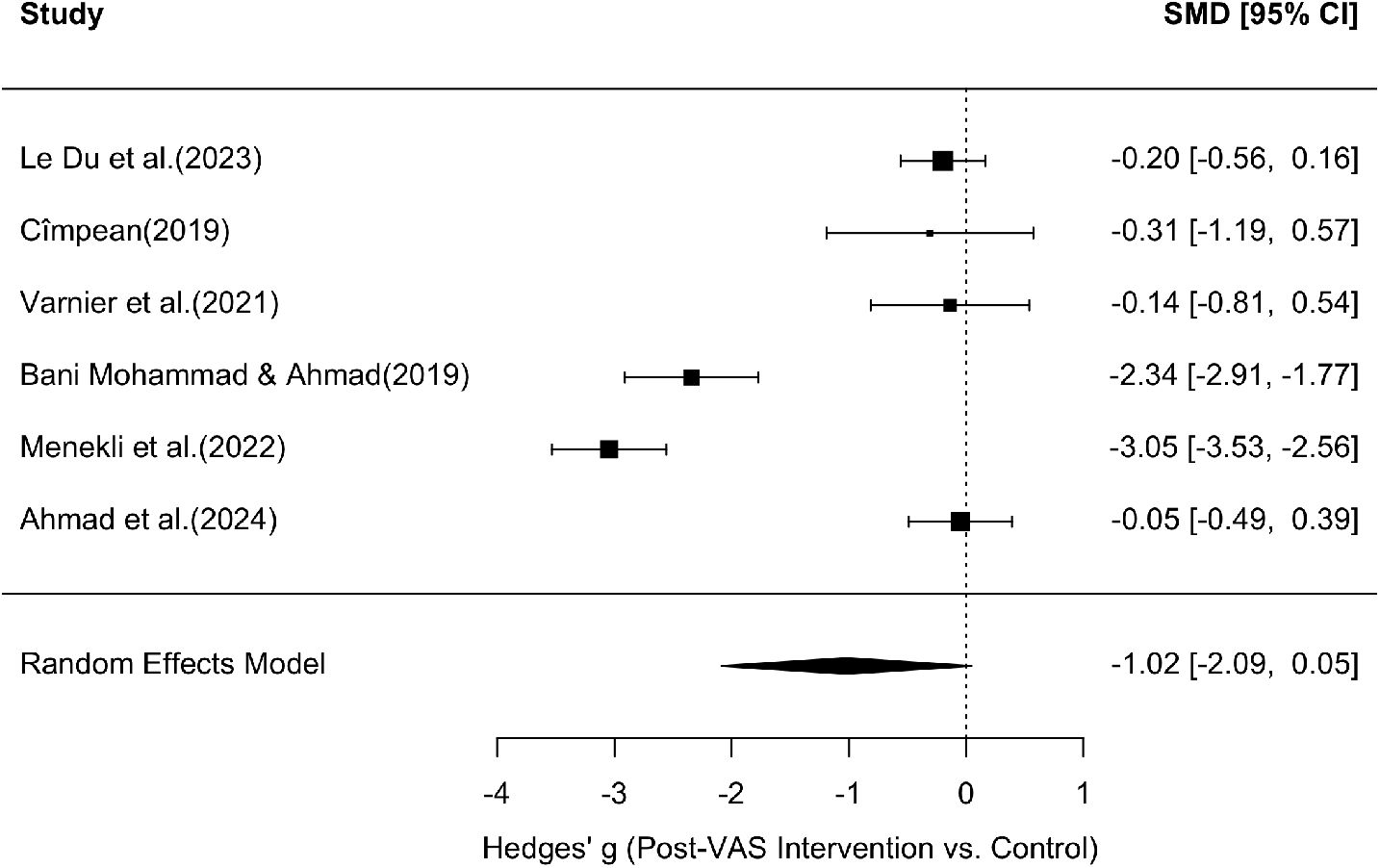
Forest plot for VAS values. Shows the results of the individual six studies and the overall result of the meta-analysis for VAS values (random effect model), as well as the respective variance.

The funnel plot (figure: 11) was visually inspected for asymmetry to assess potential publication bias. While the distribution of studies around the pooled effect size is roughly symmetrical, there is some evidence of asymmetry, particularly due to one extreme outlier (Mohammad and Ahmad, Menekli, Yaprak, and Doğan). This studies reported a much larger effect size than all others with a similar standard error, which could indicate heterogeneity rather than systematic bias.

## 5 Conclusion, Discussion and Outlook

### 5.1 Discussion

The latest developments in VR have made the technology accessible to nearly everyone. Important factors here are both the developments in hardware, such as the size and manageability of the devices, as well as the latest advances in software, which make it possible to use VR in different genres and immersion levels. This allows the devices to be better customized to the respective patients and their needs. The studies summarized here also showed many approaches aimed at a general improvement in the use of VR in cancer patients. For example, it has been shown that nature images contribute very well to general distraction and reduction of psychological symptoms such as anxiety and stress in both children and adults, or that a certain length of the application should not be exceeded to prevent cybersickness and that generally low-motion apps reduce the risk of cybersickness compared to interactive applications with a lot of movement. Children are a particularly vulnerable group, as they are very susceptible to the psychological side effects of therapy on the one hand and react particularly well to fanciful distractions on the other. Even minor examinations such as MRI or CT scans can be very stressful for children. For this reason, efforts are already being made to make such examinations as pleasant as possible for children, to allay their fears and to prepare them appropriately. For example, Liszio et al. developed a playful MRI simulation in which penguins explain the procedure to children with promising results [129]. The application is available free of charge as an app: https://www.pengunauts.com. Both the number of publications and the promising effects of VR applications in the cited studies dealing with children and adolescents show that this field is both in high demand and promising. In general, VR devices were accepted and well tolerated in all studies. Many studies also showed significant success in the treatment of psychological side effects. The success rates for physical side effects were somewhat lower, which is presumably also related to the perceived immersion of the individual test subject, which depends on many factors. VR was particularly well accepted by patients for the information provided before the respective treatment. Patients greatly appreciated visual representations of the doctor’s procedure in form of VR in all studies in this area and it reduced both fear of the application and errors made by patients during the applications. In rehabilitation after surgery, it has been shown that the use of VR in physical therapy applications helps to increase patient motivation and has therefore helped to shorten recovery times in many cases. Another interesting aspect here, however, would be the preparation for such interventions. Currently, no studies were found that deal with the preparation for interventions to this extent. VR was only used for information purposes before certain procedures. VR use in prehabilitation therefore appears to represent a research gap that requires further investigation. In surgery itself, VR has shown great potential in the use of brain mapping during brain surgery or to reduce the necessary concentrations of painkillers. In palliative care, the small, lightweight HMDs in particular have been a great benefit for the care of patients who are often bedridden and have often contributed to a significant improvement in the patient’s mood. Overall, the publications summarized here showed that VR is a promising supplement to general therapy, either to reduce side effects, to reduce the necessary administration of painkillers or other medication, or to reduce emerging care bottlenecks or long journeys for patients, for example by establishing VR support groups or therapy sessions. However, several limitations must be acknowledged. A key challenge of this review is the heterogeneity of the included studies, in terms of both study design and patient populations, which complicates the derivation of overarching conclusions. On the side of the reviewed studies, limitations include the frequent absence of homogeneous test groups, the typically short duration of VR interventions, and the limited use of control groups. To strengthen the evidence base in this field, future research should prioritize larger, well-defined study populations and extended application periods to assess the long-term effects of VR-based interventions. Moreover, the inclusion of properly matched control groups is essential to distinguish the specific benefits of VR from other therapeutic influences. Standardized protocols for VR interventions should also be developed to improve comparability between studies and facilitate metaanalyses. Addressing these aspects will help establish a more robust foundation for integrating VR into oncology care.

### 5.2 Discussion Meta-Analysis

Despite the moderate to large pooled effect size in favor of the intervention for both STAI (g = -092) and VAS scores (g = -1.02), cautious interpretation is warranted due to the lack of statistical significance. The considerable heterogeneity between studies suggests that the effect of the intervention on reducing anxiety/pain is not consistent across settings or populations. Some studies showed a significant reduction in anxiety/pain, while others showed minimal or no benefit. This variability highlights the need to consider contextual factors such as the type of intervention, timing of anxiety assessment and participant characteristics. Unfortunately, due to the heterogeneous nature of the studies, anything other than this basic analysis was not possible for an initial overview. Current evidence suggests a potentially significant effect, but further high-quality studies with standardized protocols and larger sample sizes are needed to confirm the robustness and generalizability of these results. Overall, although the results are promising, they do not yet provide conclusive support for the overall effectiveness of the interventions studied.

The findings of the risk of bias assessment further underscore the need for caution. Across both STAI and VAS outcomes, no study was rated as low risk across all domains, and the majority were judged to be at overall high risk of bias. In particular, concerns were frequent in the domains of deviations from the intended interventions, measurement of the outcome, and selection of the reported result. This suggests that methodological weaknesses, such as inadequate blinding, variability in outcome measurement, and selective reporting, may have influenced the observed results. A lack of blinding is not surprising in VR studies, as it is difficult to keep the user group unaware of their application with these devices. In addition, a general lack of control groups was a major problem in evaluating the results. While randomisation and handling of missing data were generally assessed more favorably, the predominance of highrisk ratings indicates that the evidence base is not robust. Consequently, even the moderate to large pooled effect sizes observed in the meta-analyses must be interpreted within the context of these limitations. High-quality, rigorously designed randomized controlled trials with standardized outcome reporting are needed to strengthen the reliability of conclusions in this area.

### 5.3 Conclusion and Outlook

In this systematic review, we examined the different application areas of VR in the field of oncology and categorized the different studies based on various parameters such as area of application, patient characteristics (such as age, disease and gender) and summarized the results of the different studies. The heterogeneity of the various studies and their participant groups shows the impressive diversity with which VR is finding its way into everyday clinical practice in oncology. Cancer still represents a major turning point in a patient’s life and treatments continue to have many side effects. Even if VR will never be a substitute for established standard therapies in the field of oncology, there are still some areas in which it can have a supportive effect. The psychological stress of cancer treatment, for example, often cannot be adequately absorbed in everyday clinical practice. VR can help support daily clinical practice in oncology to improve patient-centered treatment in many respects. While the current evidence suggests a potentially significant effect, further high-quality studies and, above all, standardized protocols and larger samples are needed to confirm the robustness and generalizability of these results.

## Data Availability

All data produced in the present work are contained in the manuscript

## 6 Acknowledgments

Funded by the European Union (EU) under Grant Agreement 101168715 (INSIDE:INSIGHT, https://inside-insight.eu/). Views and opinions expressed are however those of the author(s) only and do not necessarily reflect those of the European Union. Neither the European Union nor the granting authority can be held responsible for them. We further acknowledge the FWF enFaced 2.0 project (KLI 1044, https://enfaced2.ikim.nrw/), the Plattform fuer KI-Translation Essen (KITE) from the REACT-EU initiative (EFRE-0801977, https://kite.ikim.nrw/) and “NUM 2.0” (FKZ: 01KX2121). BP was funded by the Medical Faculty of the RWTH Aachen University in Germany as part of the Clinician Scientist Program. Furthermore, the authors acknowledge the Center for Virtual and Extended Reality in Medicine (ZvRM, https://zvrm.ume.de/) of the University Hospital in Essen, Germany, and the Smart Extended Reality Lab (smartXR-Lab, https://xrlab.ikim.nrw/) of the AI-guided Therapies group (AIT, https://ait.ikim.nrw/) of the Institute for Artificial Intelligence in Medicine (IKIM) in Essen, Germany.

